# Experiences of Parents and Solid Organ-transplanted Adolescents: A Qualitative Framework Analysis through Focus Groups. The TransplantKIDS Mental Health Project

**DOI:** 10.1101/2024.10.12.24315369

**Authors:** Jessica Garrido-Bolton, Rocío de la Vega, María Fe Bravo-Ortiz, Eduardo Fernández-Jiménez

## Abstract

**Background:** Solid organ transplantation is the optimal treatment for children and adolescents with end-stage organ failure. However, the biopsychosocial challenges faced jointly by the various types of solid organ recipients have received little attention from a multidimensional and qualitative approach in paediatric populations. This is the first study based on focus groups to identify the unmet care needs perceived by transplanted adolescents and parents throughout the pre-, peri-, and post-surgical phases of various types of solid organ transplantation.

**Methods:** Two focus groups were conducted with five adolescent transplant recipients and five parents, respectively, at the Children’s Hospital of La Paz University Hospital in Spain. The focus groups followed a biopsychosocial theoretical framework and used a semi-structured moderating guide. Participants were asked open-ended questions about aspects of healthcare that they would have liked to change or improve, followed by specific questions about biomedical, psychological, and social aspects that were not spontaneously addressed. The sessions were conducted using a video call, audio-recorded, and automatically transcribed. Thematic framework analysis was used to examine the data, involving familiarisation, identifying a thematic framework, indexing, charting, mapping, and interpretating.

**Results:** The findings revealed several unmet care needs, highlighting the importance of addressing social and psychological aspects of care throughout the transplantation process. Key issues common to both informant groups were inconveniences regarding the distance to the hospital and long waiting times for outpatient hospital visits, as well as the importance of peer support, humanised interaction with healthcare personnel, and mental health care during the whole process.

**Conclusions:** These findings will inform the design of future multifaceted prehabilitation interventions to prepare patients and parents for solid organ transplantation. This study highlights the importance of understanding the experiences and unmet needs of adolescent transplant recipients and their families in order to improve healthcare delivery and clinical outcomes.

## Introduction

Solid organ transplantation is the best treatment option for most children and adolescents with end-stage organ failure [1]. Despite advancements in allograft survival rates in paediatric populations following solid organ transplant, to date, understanding the challenges faced by survivors, such as the impact on biopsychosocial outcomes, has received little attention [2].

By definition, children with end-stage organ failure experience life-threatening illnesses and potentially traumatising circumstances, such as major surgeries, admission to the intensive care unit (ICU), coping with various side effects, attending regular hospital appointments, and undergoing multiple invasive testing. In addition, they experience restricted social interactions with family and friends as well as missing school and other typical normative childhood activities. Consequently, all these factors inevitably affect the quality of life of patients undergoing solid organ transplantation and their families [3].

Mental Health Consultation-Liaison teams play key roles in these clinical populations, such as assessing the suitability of living donor candidates; evaluating the recipient’s understanding of the transplant process; assessing patients’ mental state, functioning, and quality of life; treating potential psychopathological comorbidities; and delivering psychological interventions to patients and/or their relatives [4]. However, although quantitative research has contributed to these advances in the clinical assessment and treatment of organ transplant recipients, qualitative investigation is still scarce or is limited exclusively to some type of organ in paediatric populations. In this sense, this study aimed to collect and jointly analyse the subjective experiences of adolescent recipients and parents during the entire process of various types of solid organ transplantation.

The Children’s Hospital of La Paz University Hospital (Madrid) offers reference healthcare to paediatric patients throughout Spain. In particular, La Paz is the only centre in Spain that performs all types of paediatric transplants, and is a national reference centre for other complex pathologies. According to data from the National Transplant Organization (ONT, acronym in Spanish) in its annual report, La Paz is the Spanish centre that performed the highest number of paediatric transplants in Spain in 2022, making it a suitable context for collecting clinically relevant data in these paediatric populations [5].

To the best of our knowledge, this study is the first to qualitatively identify unmet care needs, utilising focus groups, of paediatric patients and parents throughout the pre-, peri-, and post-surgical phases of diverse solid organ transplants. This qualitative approach contributes to the humanisation of scientific and care activities as it incorporates and integrates the perspectives, preferences, values, and expectations of the recipients of the care process. In this sense, this study follows a family-centred care model [6] and promotes a comprehensive and holistic care process in which patients and their parents are represented, thereby humanising health care by obtaining clinical demands directly from primary stakeholders.

We expect that these findings will provide essential novel information for the design of future prehabilitation interventions for paediatric transplants (i.e. to prepare the patient to be in the best possible state before surgery).

## Materials and Methods

### Study Design

The focus group method was selected because of its group dynamics, which can generate rich data, particularly in situations where prior knowledge is limited [7]. Focus groups and semi-structured interviews are widely recognised as the primary methods used in qualitative research [8]. Non-quantitative methods have been increasingly used by paediatric psychologists and other researchers to improve the understanding of the scientific community and provide new perspectives in healthcare [9]. These methods examine how children and their families experience and respond to health-related challenges, allowing researchers to better understand the phenomenon under study [10].

To ensure the accuracy and transparency of the reporting of this qualitative study, we closely followed the COnsolidated criteria for REporting Qualitative research (COREQ). This involved a 32-item checklist developed specifically for reporting research related to interviews and focus groups [8].

### Sample

Focus group participants were recruited by the Mental Health Consultation-Liaison team of the Children’s Hospital of La Paz University Hospital (Madrid, Spain), which approached eligible candidates. Suitable candidates for the patient group were adolescents between the ages of 12-18 who had already undergone solid organ transplantation and were able to speak Spanish fluently. To enrich the content of the focus group and ensure greater diversity of experiences, participants who had undergone transplantation of different solid organs were selected deliberately. The patient group consisted of adolescents aged between 13 and 18 years (mean = 15.6; *SD* = 1.95 years) who had undergone transplantation, on average, 13.4 months ago (*SD* = 6.54; range: 6–24 months).

As in the adolescent group, parents of children who had received different solid organ transplants were prioritised in the parents’ group. The parents chosen were not the parents of the adolescents in the focus group to broaden the spectrum of experiences and perspectives. However, an exception was made for one adolescent who received a lung transplant due to the much smaller sample size of this type of transplant, thus allowing for the joint participation of the mother and child. The second group consisted of parents of patients aged between 8 and 15 years (mean = 11.6; *SD* = 3.05 years) who had undergone transplantation, on average, 14.6 months ago (*SD* = 5.46; range: 10 to 24 months).

In the adolescent group, we included more than one participant who had received a kidney transplant, and in the parent group, we included more than one parent of a heart transplant recipient, as we considered these types of transplants to be the most representative. The characteristics of the participants are listed in Table 1.

**Table 1.** Characteristics of study participants (*n* = 10).

| **Participants:** |  |  |
| --- | --- | --- |
| **Adolescent Group** | **Sex** | **Type of transplant received** |
| Adolescent 1 | Female | Kidney (deceased donor) |
| Adolescent 2 | Female | Heart |
| Adolescent 3 | Female | Kidney (deceased donor) |
| Adolescent 4 | Male | Lung |
| Adolescent 5 | Male | Liver |
| **Participants: Parent Group** |  |  |
|  | **Sex** | **Type of transplant their child received** |
| Parent 1 | Male | Heart |
| Parent 2 | Female | Heart |
| Parent 3 | Female | Kidney (living donor) |
| Parent 4 | Male | Hepato-renal |
| Parent 5 | Female | Lung |

### Procedure

The parents of each enlisted participant (or, when applicable, patients of legal age) were contacted by telephone by one of the team researchers to ensure that the participant met the criteria and to explain how the group would be conducted. All the contacted participants wished to participate in the study. Parents of patients (or, when applicable, patients of legal age) who provided written informed consent were scheduled to participate in this study. One person did not show up to the adolescent group and was contacted later for an individual in-depth interview following the same format and conditions.

The patients’ parents were asked to authorise, in an informed manner, the participation of their children and themselves (when required) in focus group sessions to qualitatively identify unmet care needs during the pre-, peri-, and post-transplant phases. Two group sessions were conducted (one session per participant type: patients and parents), lasting approximately 90 min each, arriving at data saturation (meaning that no new value information was obtained). The groups comprised five participants each (*n* = 10 in total).

The interview guide used in both focus groups followed a biopsychosocial theoretical framework. Specifically, each focus group began indicating group norms (confidentiality/privacy, not interrupting others’ turn to speak, and not judging others’ comments). Afterwards, the session continued with a first part of open questions about what aspects participants would have changed or improved in health care, both for the patients and parents, during the entire transplantation process. Subsequently, the second part of the focus group posed specific questions about the biomedical, psychological, and social aspects that would have improved throughout the transplantation process and were not spontaneously addressed in the first part of each focus group. The facilitator followed a semi-structured moderating guide, and the questions posed are listed in Table 2.

**Table 2.** Focus group topics and key questions.

| Adolescent Focus Group |  |
| --- | --- |
| Free open-ended questions phase | - What aspects would you improve during your visits to the hospital or during your admission to the hospital for the organ transplant you received? |
|  | - Have you missed any kind of support or specific help related to the transplant from any kind of professional working at the hospital? |
| **Directed open-ended questions phase\*** | - In terms of the care you have been given by the **doctors and nurses**, what (other) aspects would you improve during your visits to the hospital or during your admission to the hospital for the organ transplant you received?<br>- In terms of **psychological and emotional issues**, what (other) aspects would you improve during your hospital visits or during your admission to the hospital for the organ transplant you received?<br>- In terms of **social support**, what (other) aspects would you improve during your visits to the hospital or during your admission to the hospital for the organ transplant you received? |
| **Parent Focus Group** |  |
| **Free open-ended questions phase** | - What aspects of your child's health care during the organ transplantation process that he/she has undergone, either during the follow-up in outpatient appointments or during hospitalisation, would you improve?<br>- What kind of interventions or clinical care have you missed during this process for your child?<br>- What kind of interventions or clinical care have you missed during this whole process for you?<br>- What aspects of health care would you improve for you as a parent of a child who has undergone an organ transplant? |
| **Directed open-ended questions phase\*** | - On a **medical level**, what (other) aspects would you improve in the care of your child and you as parents during the solid organ transplantation process?<br>-On a **mental health level**, what (other) aspects of care for your child and for you as parents during the solid organ transplantation process would you improve?<br>-On a **social level**, what (other) aspects of the care for your child and for you as parents during the solid organ transplantation process would you improve? |

*\*Directed questions were asked only when participants did not spontaneously address specific aspects of one of the three areas of the biopsychosocial model.*

The sessions were conducted by videocall, audio-recorded, and automatically transcribed, respecting the confidentiality of the information at all times. The sessions were facilitated by a member of the research team and not by the PI of this project (EFJ) to avoid bias (the PI knows most of the patients and parents). The facilitator was a female psychologist with a Master’s degree. However, the facilitator was introduced as a researcher and not as a psychologist, so as not to bias participants in prioritising mental health aspects to the detriment of biomedical and social domains. In addition, this facilitator worked mainly on another PI’s scientific project, focused on a different paediatric clinical population, and did not have any relationship with the participants prior to study commencement, nor did they know of her previously. Furthermore, this facilitator did not participate in any of the subsequent stages of this project: transcription generation, data analysis, data interpretation, writing or reviewing this paper.

Subsequently, the automatically generated transcriptions were reviewed to (1) change the name of the participants using a code and (2) correct any transcription mistakes. The recordings were maintained until it was verified that the transcription process had been performed correctly. The transcripts were not returned to participants for their comments. When needed, in anticipation that more relevant information would be collected, participants were offered the possibility of conducting individual in-depth interviews after the focus group. Interviews were conducted using the same standards mentioned above.

The study protocol was registered at clinicaltrials.gov on 1 July 2022 (NCT05441436) [11]. Recruitment for this study began on October 21, 2022 and ended on February 8, 2024.

### Ethical Considerations

This study was conducted in accordance with the principles established in the Helsinki declaration, strictly respecting confidentiality and the requirements of Spanish (14/2007, 3 July 2007 on Biomedical Research; and Organic Law 3/2018, 5 December 2018) and European data protection regulations. This study was approved by the Research Ethics Committee of La Paz University Hospital (Madrid, Spain) (institutional code: PI-5223). Parents or legal guardians of patients (or, when applicable, patients of legal age) provided their written informed consent, for minor patients and for themselves, to participate in this study.

### Data Analysis

Two coders (JGB and EFJ) independently analysed the data following the steps of the “qualitative framework analysis” [8]. The first step was familiarisation, which involved reading all the transcripts thoroughly, listening to the tapes, and listing key ideas/recurrent themes to identify a thematic framework (immersion in data). A coding frame was developed based on these key themes. This involved entering brief summaries into the coding frame, along with the page numbers of the original data excerpts, facilitating easy retrieval for further analysis.

Subsequently, a biopsychosocial model was systematically applied to all the data (indexing), and the data were rearranged according to the relevant sections of the thematic framework (charting). Charting was performed using columns (participants) and rows (themes). The coders had a meeting with a third researcher with experience in framework analysis (RV) to resolve any coding disagreements. Finally, concepts were defined, and data were interpreted by comparing and contrasting respondents’ accounts and searching for explanations of patterns observed in the data (mapping and interpretation) [9].

## Results

In this section, we present participants’ experiences regarding solid organ transplantation and biopsychological factors resulting from the analysis of the transcripts. Table 3 presents an overview of the topics that emerged during the focus group sessions.

**Table 3.**
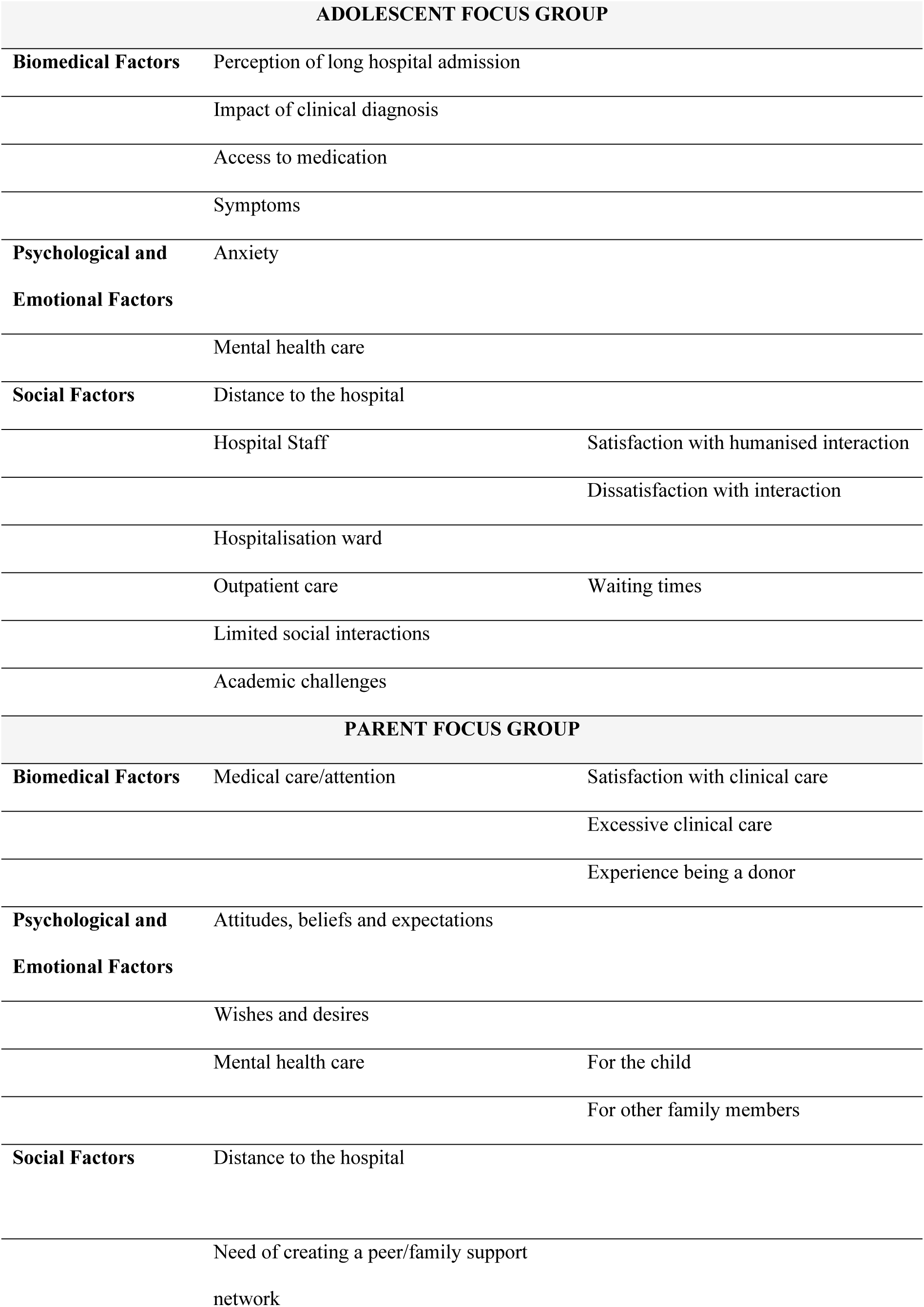
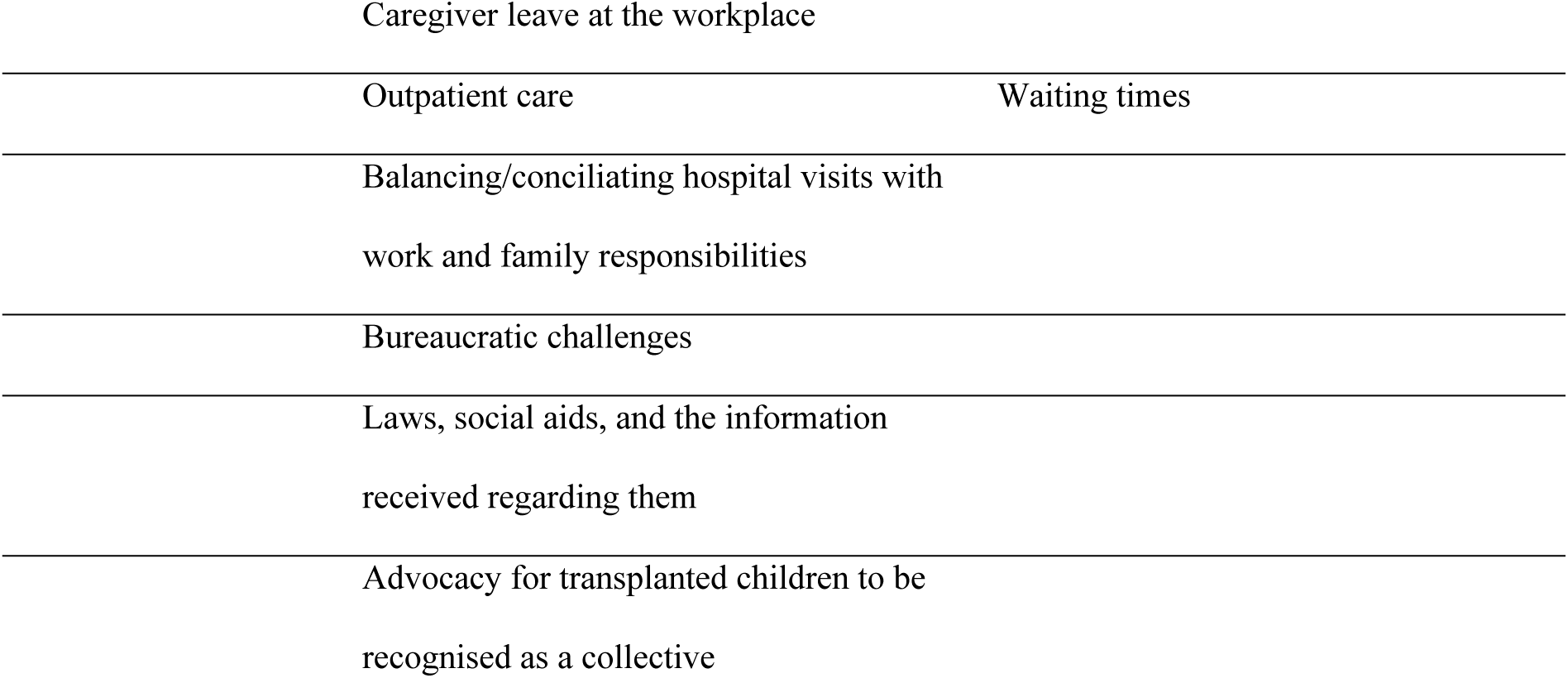
Key issues addressed by each focus group: adolescents and parents.

### Adolescent Focus Group

#### Biomedical Factors

##### Perception of long hospital admission

Most patients felt that their hospital stay was excessively prolonged. One individual mentioned that it became particularly burdensome once she experienced clinical improvement.

> *“I felt it was overwhelming to be hospitalized for a very long time”.*

> *“When I started to get better, I got tired of being there [hospital ward] because I was there for a long time”.*

##### Impact of clinical diagnosis

One adolescent requiring a kidney transplant, described the impact of suddenly and unexpectedly receiving the diagnosis of a chronic renal condition.

> *“Before the transplant, I was diagnosed with renal failure from one day to the next, and I had no symptoms or anything… So, I was very shocked by that.”*

##### Access to medication

Certain individuals mentioned that access to medication could be problematic, as it was intermittently unavailable at local pharmacies.

> *“Sometimes health professionals had trouble getting me certain medications because they were either out of stock or temporarily discontinued”.*

##### Symptoms

The most common symptoms were fatigue, sickness, and concentration difficulties.

> *“I felt very weak and tired at the hospital and at home, I didn’t want to get out of bed… I felt a lot of stress”.*

> *“My immune system was very low and it made it [studying] more difficult (…). I could not concentrate, grasp the information well, understand what the teachers were saying…”.*

#### Psychological and Emotional Factors

##### Anxiety

Most participants reported feeling anxious as their predominant emotional state. One adolescent who became a kidney recipient, reported frequent panic attacks following renal graft rejection.

> *“To be honest, I experienced a lot of anxiety after the transplant, especially at the hospital”.*

> *“I wish the transplant had gone better… I had a lot of panic attacks in the ICU because of my rejection.”*

##### Mental health care

Adolescents rated contact with a mental health professional (clinical psychologist/psychiatrist) as a positive intervention. One adolescent who received a lung transplant, noted that he benefited from psychological assistance to cope with medical tests that caused him anticipatory anxiety.

> *“I value psychologists the most. Before my transplant, I was anxious and stressed. The psychologists helped calm my nerves during that time, because just thinking about the transplant procedure made me anxious.”*

Another adolescent who underwent a liver transplant, commented: *“The psychologists had to intervene because I was clearly reluctant to undergo the lumbar puncture”*.

#### Social Factors

##### Distance to the hospital

Adolescents who had to travel between cities for medical attention reported some inconveniences, such as transportation and being far from their loved ones.

> *“Transport to medical appointments, that’s the only problem (…) the type of vehicle [ambulance] was uncomfortable …”.*

> *“Personally, I strongly would have preferred being in my homeland [region of Spain], I missed having friends visit [during hospital admission] because I was in Madrid.”*

##### Hospital Staff. Satisfaction with humanised interaction

Overall, there was widespread satisfaction with the interpersonal care provided by the hospital staff among the participants, which significantly eased their hospital visits and admissions.

> *“All the efforts made by the nurses, doctors, and orderlies to create a positive experience for us despite the challenges (…). Their efforts ultimately gave me hope for life after the transplant and, at the same time, made my time there a little brighter.”*

Only one adolescent held a more critical view of this matter.

> *“I think I would improve the interaction from the nurses or assistants (…). Sometimes, they would come in with a bit of a bad mood or something like that. That’s the only thing I would change.”*

##### Hospitalisation ward

One adolescent who was a kidney recipient, expressed discontent with the formal aspects of the hospitalisation ward, particularly the distribution of patients in the rooms.

> *“In other surgeries, such as the catheter dialysis (…) I was in a shared room with a two-month-old baby. Therefore, the truth is that I had a very, very bad time (…). I told my doctors “I want to leave now because I cannot stay here any longer”. Because imagine, a two-month-old baby girl who would not stop crying all night long.”*

##### Outpatient care

Adolescents expressed complaints about their waiting times.

> *“I had to wait for two or three hours for some appointments (…). When I had to go there every three months, I was like – I do not want to go because I am going to be there for hours.”*

##### Limited social interaction

All the adolescents agreed that this was one of the biggest challenges.

> *“When I was on dialysis, I had very limited freedom. Travelling, for instance, was quite challenging because of the machine and logistical issues involved. It restricted me during a time when my friends were enjoying more independence and going out at 16-17 years old. This was particularly tough for me.”*

##### Academic Challenges

All adolescents, particularly those in their final years of high school, expressed the negative impact of transplant on their academic performance.

> *“Truth be told, I had a really tough time because I missed a lot more school than usual (four and a half months), and I was really stressed out because I was at risk of repeating a year, which isn’t ideal during high school”.*

> *“When you return to high school after a transplant, it can be in the middle of the academic year… it can be disorienting because your classmates are further ahead than you”. Many expressed frustration and despair over losing valuable school time due to the long waiting hours for outpatient hospital appointments.*

> *“The truth is that I was in my first year of high school and waiting two or three hours for medical check-ups wasted a lot of my time”.*

One adolescent expressed dissatisfaction with the lack of curricular adaptation.

> *“When I arrived, I had missed most of the first term, and unfortunately, the teachers didn’t offer much assistance. They immediately gave me exams without allowing me time to catch up, which caused me to struggle. Now, I find it challenging to keep up with the others.”*

Finally, some adolescents expressed limitations in receiving educational support either at home or at the hospital, indicating that their assistance was insufficient. One adolescent, who was entitled to the Home Education Support Service, reported not having benefited from this service at all.

> *“They told me that since I was in my final school year I could request home education for when I had to stay home in isolation, to help me keep up and not fall behind. However, the teacher never showed up (…). It really frustrated me, as it has been very noticeable, especially in mathematics which is a subject where a lot of progress is made”.*

> *“When I was in the hospital, there were two teachers who came to help (…), but I could see that they didn’t have enough time because there were too many children, too many people…”.*

### Parent Focus Group

#### Biomedical Factors

##### Medical care/attention

Overall, most parents expressed a high level of satisfaction and gratitude for their children’s medical follow-up and care throughout the transplantation process.

> *“We are very happy and grateful with the medical attention our child received. The truth is that the doctors were always excellent.”*

> *“It seemed very human to us that when our son was undergoing the operation, the surgeons came out to explain what was happening. You are waiting there for many hours, and the fact that they came out to provide us with updates… It seemed to us that we always received information; they always asked you how you were doing and so on.”*

However, one parent reported a subjective experience in which, on some occasions, medical attention seemed excessive.

> *“There are times when you get overwhelmed by all the medical attention. When you think about it rationally, you realise it is good that they are so thorough in attending to him and monitoring his condition. They want to ensure that everything is under control, and that they are doing well. It’s just that we spend all day at the hospital, and at first glance, it can feel excessive.”*

One woman expressed dissatisfaction with the medical attention she received after donating a kidney to her son.

> *“In my particular case, I was the donor, and naturally, all the attention was focused on the child, not the parent. However, I had also undergone surgery… In some way, I felt neglected, receiving minimal attention as an adult. This lack of care was concerning, setting aside a more emotional aspect, I could have experienced a medical complication.”*

#### Psychological and Emotional Factors

##### Experiences, attitudes, beliefs, and expectations

Parents shared their experiences and thoughts when undergoing their children’s transplantation process.

> *“There were good days and bad days, but I’ve been very nervous and upset. I did not know what was going to happen, and everything could change in five minutes (…). The worst time for me was the month after the transplant when my daughter was admitted to the ICU. I only noticed the shortcomings of nurses, assistants, and doctors. I just wanted everyone to be with my daughter. I did not want to accept that it was my daughter’s body that was sick; instead, I tended to blame others. Now that I am out of that rollercoaster, I look back and can understand it was very difficult, very complicated, and can see that everyone did their best. I am very grateful now and I have come to realise it was my nerves, the tension, the fear….”*

> *“I felt a lot of fear, I wanted someone to tell me exactly what a heart transplant was like, what could happen… As time went on and I met more families, I came to understand that the anxiety I had about knowing what would happen couldn’t be eased by another parent. I realised that my daughter’s transplant would be unique to her. They might have performed fifty transplants before, but I understood that when they transplanted her, she would face her own set of complications, just as other children would face theirs.”*

##### Wishes and desires

We asked parents if there was anything they wished they could have had or felt they missed during their child’s transplant process. One parent expressed that they missed the opportunity to convey gratitude to their son’s donor.

> *“There is one part that has always stayed with me, like a thorn in my side. I think it may be dictated by protocols that it cannot be done, but I always feel a debt of gratitude to the donors. It’s like you receive the donation, and have not been able to thank them for anything. It is something that makes me feel like I have a debt I cannot repay. I imagine that it probably cannot be done, and so on. But emotionally, if there is one thing I still need to do, it is to thank the donor.“*

##### Mental health care

Parents discussed which stages of the transplant process they believed psychological support was most useful or necessary for their children. Several parents noted that their children began expressing sudden concerns about death in the period leading up to transplantation.

> *“When the idea of the transplant was introduced, she did need psychological treatment. The treatment was not very, very demanding, but we did notice that from then on, she needed it, as she had changed a little bit. She started to be quieter, she started to think more… Anyway, she started asking us things about death… Now, we still go to the psychologist, but she is doing very well.”*

> *“In the months before the transplant, I noticed my son was very worried about death and related issues. I do not know if it’s a normal concern or not at that age, but I think psychological care beforehand could have helped.”*

Another fundamental aspect discussed was the necessity of providing psychological support not only for the child undergoing the transplant but also for other family members, such as siblings, who also experience distress during this period and may feel overlooked.

> *“My eldest daughter had a very bad time and kept quiet about it. I think siblings are also victims in a way, and we only realise this later when everything calms down. When the calm comes after the tsunami, then you start to notice the other little people around you, reminding you that they exist and that the situation has also affected them (…). Psychological care is provided to the parents, but in my case, the siblings are somewhat left in no man’s land (…). They are left alone at home with their grandparents or other family members and are often overlooked as the days go by.”*

#### Social Factors

##### Distance to the hospital

Parents expressed the burden of not being able to receive medical attention in their own city, which forced them to commute to the city where the hospital is located. This situation disrupted their normal lives and resulted in significant economic loss.

> *“Since we are from far away [a Spanish region far from the city where the hospital is located], we had to move and stay in Madrid for the whole time we were in the hospital and for the post-transplant period, as you have to go up to four times a week or several times a day to the hospital. These trips to Madrid have a cost and they do not allow you to make up with overtime at work.”*

> *“If you live near [the hospital] that’s fine, you have it easier, but those who do not live near, have to spend a lot of money on travel, for example, on lodging… and that, in the end, of course, is a stress, because you say gee… Can I do it? Can I afford it? I think that’s a big problem.”*

Finally, some expressed how difficult it was to be far from their loved ones and unable to leave the city while their daughter was on the waitlist for heart transplants.

> *“We were on the waitlist for three years. We have lived in Madrid for nine years since my daughter was born, but our family is in Granada [a region of Spain]. It was a tough three years because we could not leave Madrid.”*

##### Need of creating a peer/family support network

Parents agreed that it would have been beneficial for them and their children if the hospital had offered a peer/family support network, allowing them to meet other children and families in similar situations.

> *“It would be good for the children, especially in a medical specialty such as lung transplantation, which is not very common as there are few cases and few children who have gone through this process and are alive; to be able to do some kind of activity for them to interact more, get to know each other, share their experiences. Because in my son’s case, he is now entering adolescence and just two weeks ago, he met another boy who had also received a lung transplant, and for him, it was something cool – he has gone through the same thing as me – he said. They need to share that information, they need to hear the opinion of someone else who went through the same process.”*

> *“Those three years [waiting for her daughter’s heart transplant] I would have liked to have met more parents in the same situation”.*

##### Caregiver leave at the workplace

One parent, although grateful for the flexibility offered at work, wished they had known what the law states about time off in such situations.

> *“At work, they told me to take as many days off as necessary, but I thought maybe in other places there are more limited days of time off, or there is an established protocol… Because especially after the transplant you have to be at the hospital for a long time and I think it is more than the legal time allowed”.*

##### Outpatient care

Some parents expressed complaints about waiting times.

> *“I don’t know what the hospital protocol is like or anything, but you arrive with a child who has been very ill and still is, and you have to wait three hours for an appointment… I do not understand why they do not give them a card or something to help respect the appointment times. If they had to wait forty minutes, okay, but what is not acceptable is three or four hours. And if you have another appointment that day, it means that for two consultations, you could be losing up to five or six hours.”*

Concerns were also voiced about remaining in a crowded waiting room with an ill child who was exposed to many other children for extended periods.

> *“When you’re in a waiting room with forty other people and you have to stay there for three or four hours, you are putting your daughter at risk of catching anything. They are children who have been transplanted and well, my daughter was also weak before she was transplanted, so, I think that should be improved.”*

##### Balancing/conciliating hospital visits with work and family responsibilities

This aspect was a shared concern among the participants of the group.

> *“There is significant stress for us as parents trying to be in two places at once – work and the hospital. The challenge is even harder if you have more children. We need to make a living but it’s hard to balance the fact that you have to dedicate yourself to your son who is in post-operative care, at the same time that you are leaving work and family behind.”*

> *“In my case, we spent six months in hospital, from the time she was transplanted until she came home. A long time passed and my other children felt in some way abandoned. – Where are my parents? – they must have thought. And of course, you get home, and you need to sleep because you are knackered, and all you really want to do is go back to the hospital because my only obsession was to be in the hospital. My eldest daughter had a bad time because, of course, I could not look after her, my husband could not look after her…We spent most of our time at the hospital.”*

##### Bureaucratic challenges

There was widespread discontent with how time-consuming and inefficient bureaucracy can be, especially when you have a child who is ill and needs full attention.

> *“Paperwork is very slow and time-consuming in general, and even more so for those of us who are parents of children who are ill and need all our time, they need us to be with them 100%. We do not have time to be doing all these procedures and going from one place to the other because the children need 100% of the parents’ care.”*

> *“You have to navigate through a lot of bureaucracy. For instance, with my daughter, they approved her dependency status because before her operation, she had only half a heart and needed oxygen. However, it took two and a half years to get that approval. Another frustrating issue is how they assess her disability level; it can change if her condition improves or after surgery, and then you are back to fighting bureaucracy again, all while caring for a sick child at home. It is unpredictable—today they grant you assistance, but tomorrow they might revoke it. For instance, after a successful transplant, she might need a tracheotomy, so the situation keeps changing, and you cannot constantly update the administration about the evolving circumstances.”*

##### Laws, rights aid, and the information received regarding them

Some parents were disappointed to discover very late, or not at all, certain laws that could have provided crucial support during their child’s transplant.

> *“Nobody informed us that there was a law that covered parents with children with serious illnesses and allowed you to reduce your working hours as from 50% up until 100% (…). I found out very late that I could have dedicated myself exclusively to the care of my son and still receive my full salary.”*

They also missed having a reference figure to whom they could direct their questions about laws and available social aid.

> *“I missed having a specific figure to inform us about all the regulations, laws, aid, and procedures necessary when you have a child with special needs and requires special attention. There are likely many laws or regulations, whether at the State or community level, that I am unaware of and may only learn about in the future. I miss having a unified figure to guide us. Today, I feel we are excluded from all the help offered by the Country and the community because we simply do not know about it. There is no one to inform us.”*

One woman described feeling misinformed and struggling to understand the processes because she was an immigrant and unfamiliar with many laws and regulations.

> *“I don’t know the policies, I don’t know the Country’s system… I do not know how to get an appointment to be informed… It has been very hard for me. I have been doing this for three years now, and I do not see that there is an organisation offering help to a parent who does not know or is not from this country. I do not have the degree of Dependency…I have knocked on the doors of the social workers and I have not managed anything.”*

A woman expressed disappointment that psychological support provided by national insurance is not available in her homeland for children who have undergone transplants.

> *“Here in Gran Canaria, psychological support for a transplanted child isn’t considered necessary, so we had to pay for private consultations. Unlike getting a prescription for medication, psychological treatment takes time, and sometimes the financial burden is too much to bear. We were told by the national insurance that unless there is something like Attention-Deficit/Hyperactivity Disorder (ADHD), or a condition of the autistic spectrum or a dysfunctional family, that my son was not eligible for psychological therapy.”*

##### Parents’ advocacy for transplanted children to be recognised as a collective

> *“My son faces both nephrological and visual challenges. The main issue is that transplanted children are not recognised as a distinct group. For instance, my son receives comprehensive support for his visual impairment from the Department of Education, including a social worker, teachers, and adapted materials, because individuals with visual impairments are recognised as a group. In contrast, transplanted children do not receive similar support— there are no counsellors or social workers available to us. For my son’s visual impairment, we have regular visits, phone calls, appointments, and guidance on aids. However, for his transplant-related needs, there is minimal support and recognition. It is crucial for transplanted children to be acknowledged as a distinct collective group, similar to how other groups such as those with visual impairments or mobility issues are recognised and supported.”*

## Discussion

To the best of our knowledge, this is the first study to qualitatively examine the perceived unmet needs throughout the paediatric organ transplantation process, comparing the subjective perspectives of both adolescent patients and parents regarding several biopsychosocial domains and jointly covering various types of solid organ transplants. This work revealed significant insights into the experiences and perceptions of adolescents and parents concerning medical care, social dynamics, and psychological challenges associated with undergoing a solid organ transplant. We now discuss the findings identified as most relevant from both focus groups and compare these results with the existing body of literature. Furthermore, we conducted a comparative analysis of topics that emerged across the two focus groups.

On one hand, a significant finding was the disparity between the focus of parents and teenagers on academic concerns. In particular, parents tended not to mention academics, whereas adolescents frequently expressed overwhelming anxiety regarding their academic performance, which was exacerbated by a perceived lack of educational support and adaptations. Additionally, existing literature suggests that adolescent transplant recipients may exhibit cognitive and neuropsychological impairments [12,13], which may contribute to the aforementioned perceived problems. This underscores the need for more robust academic support systems tailored to adolescents’ unique circumstances [14].

On the other hand, key issues that emerged for both informants were inconveniences regarding the distance to the hospital, long waiting times for outpatient hospital visits, the need for peer support, satisfaction with the humanised care provided by healthcare workers, and the relevance of mental health support during the transplant process.

In particular, the experience of Adolescent 5 underscores a common challenge faced by patients who must travel to another city for transplant procedures because they reside in other regions. In this sense, La Paz University Hospital is a national reference hospital in Spain and is the only hospital to date that performs all paediatric transplants. The logistical complexities of long-distance travel, often via ambulances or planes, add a layer of stress to the already taxing situation. Moreover, distance to the hospital imposes an economic burden on families, who must cover the costs of lodging and related expenses. Being far from loved ones during difficult times can also be emotionally distressing. Peretz et al. [15] examined the psychological and financial challenges associated with long-distance travel for liver transplantation. While patients generally remain confident in their healthcare providers and do not feel that their care is compromised by distance, many, particularly women, expressed dissatisfaction with the limited opportunities to support visits. Additionally, the economic burden of being far from home likely contributed to 70% of patients preferring to undergo the procedure locally. These factors suggest a need for more localised transplant services or improved support for travel and lodging logistics to reduce the burden on patients and their families.

In addition, extended outpatient waiting times were generally poorly tolerated, and prolonged hospital stays were particularly challenging for the adolescent group. This highlights an area that requires urgent improvement to enhance patient and caregiver satisfaction, as previously noted in the literature on paediatric outpatient care [16].

Furthermore, both parents and adolescents identified social issues as their predominant concern over biomedical and psychological issues. This finding is consistent with the current research, which shows that patients and families face enduring psychosocial risks long after the initial post-transplant phase [17]. Consequently, it is essential to make social support a central component of interventions to address the well-being of patients and their families effectively.

Among the topics that emerged at a social level, participants conveyed that they would have greatly valued the opportunity to engage in peer and family support groups. In particular, the adolescent group in this study expressed being particularly affected by periods of limited social interaction due to hospital admissions and necessary isolation protocols. Social isolation can be distressing, emphasising the importance of creating robust communication channels and support systems to mitigate the impact of isolation on patients’ mental health [18]. This is consistent with the work of Anthony et al. [19], who emphasised the significant benefits that adolescents gain from peer support programs, reporting the satisfaction perceived among participants in an online mentorship program designed for paediatric transplant patients. Such peer groups provide critical emotional support and a sense of community, which can significantly improve the coping mechanisms of both patients and their families.

Furthermore, one of the notable social-level suggestions highlighted by participants in the parent group was the potential benefit of having a unified reference figure from Social Work to assist with bureaucratic procedures systematically rather than reactively. This would alleviate the administrative burden on families, allowing them to focus on emotional and psychological aspects of care. There was a strong desire for a dedicated reference person who could provide expert guidance on complex laws and regulations related to medical care, economic aid, and work-leave entitlements. For instance, in Spain, Compensation for Leave to Care for Children with Cancer or Serious Illnesses (CUME, acronym in Spanish) [20] allows working parents to take leave to care for children undergoing serious medical treatment, with financial support provided during their absence. However, one parent noted that they only learned about this social aid during the focus group discussion of this project and wished that they had been informed earlier.

The difficulty of balancing family responsibilities with work commitments has also been highlighted as a significant challenge. This difficulty aligns with the existing literature, which categorises the challenges faced by parents of children with chronic illnesses into three domains: work, family, and personal. Workplace challenges commonly include securing flexible work arrangements, managing absenteeism, and encountering unsupportive employer attitudes. Family-related issues involve finding suitable childcare facilities, managing time constraints, supporting ill children, and fulfilling overall family duties. On a personal level, parents frequently experience stress, uncertainty about the future, and the feeling of “having to do it all” [21]. This calls for more flexible support systems to assist families in effectively managing their dual responsibilities.

Additionally, addressing the needs of siblings was another critical point. Existing literature indicates that siblings of children with chronic conditions face unique challenges that affect their health-related quality of life, including negative emotions, separation anxiety, increased responsibilities, and feelings of exclusion [22]. Feelings of abandonment among siblings must be mitigated by including them in the entire care process and offering psychological support, as necessary. This holistic approach ensures that the emotional needs of the family are met, thereby promoting a more cohesive support system.

It is possible that all of these perceived needs could be partially addressed by the suggestions made by one of the parents during the focus group. They proposed that children undergoing transplantation should be legally categorised as a distinct group regarding disability recognition. This recognition would streamline access to the necessary support services for these children and ensure that parents receive accurate and timely information.

Finally, humanised interaction by healthcare personnel was highly valued by both patients and parents, underscoring the importance of empathy and compassion in medical care [23]. Additionally, both groups agreed that mental health support was beneficial throughout the transplantation process. In particular, this study highlights the pressing need to address thoughts of death and the associated psychological impact among these patients. Consequently, it is imperative to improve mental health interventions to provide comprehensive support. Accordingly, the mental health profiles of diverse paediatric organ transplant recipients are being increasingly investigated to ensure the provision of holistic care for these clinical populations [2,11,24].

### Strengths and Limitations

Regarding the limitations of this study, it included a modest sample size from a single hospital. Nevertheless, the participants were selected purposively to encompass a representative view of the diverse potential circumstances surrounding the paediatric transplant process in accordance with sociodemographic and clinical criteria (organ type, sex, and socioeconomic status). Furthermore, despite the potential for conformity processes among participants in the focus group methodology, members in both groups conducted for this study expressed divergent perspectives and nuanced opinions, demonstrating their willingness to articulate discrepancies with other participants.

### Conclusions

In conclusion, despite these limitations, this study suggests several key areas for improvement in the care and support of adolescent transplant patients and their families. Enhancing academic support, reducing outpatient waiting times, the humanised interaction by healthcare personnel, providing mental health and comprehensive family support are critical steps towards improving the overall care experience and represent key areas for future research. Additionally, the establishment of a unified Social Work figure to assist bureaucratic processes is also considered essential in addressing the multifaceted needs of these patients and their families. These findings will inform the design of future multifaceted prehabilitation interventions for the entire paediatric transplant process.

## Data Availability

All relevant data are within the manuscript and its Supporting Information files.

## Acknowledgments

The authors acknowledge Dr Antonio Pérez-Martínez’s contribution in funding acquisition for this project. Furthermore, the authors extend their appreciation to all healthcare professionals who referred patients to this study. Finally, the authors express their gratitude to the psychologist who assisted the research team in conducting group sessions and in-depth interviews.

## Notes

### Competing Interest Statement

The authors have declared no competing interest.

### Clinical Protocols

https://www.frontiersin.org/journals/psychology/articles/10.3389/fpsyg.2024.1308418/full

https://clinicaltrials.gov/study/NCT05441436

### Funding Statement

Yes

### Author Declarations

This study was conducted in accordance with the principles established in the Helsinki declaration, strictly respecting confidentiality and the requirements of Spanish (14/2007, 3 July 2007 on Biomedical Research and Organic Law 3/2018, 5 December 2018) and European data protection regulations. This study was approved by the Research Ethics Committee of La Paz University Hospital (Madrid, Spain) (institutional code: PI-5223). Legal guardians of patients (or, when applicable, patients of legal age) provided their written informed consent to participate in this study.

